# Perceptions of barriers and facilitators for cervical cancer screening from women and healthcare workers in Ghana: Applying the Dynamic Sustainability Framework

**DOI:** 10.1101/2024.02.22.24303192

**Authors:** Adwoa Bemah Boamah Mensah, Thomas Okpoti Konney, Ernest Adankwah, John Amuasi, Madalyn Nones, Joshua Okyere, Kwame Ofori Boadu, Felicia Maame Efua Eduah, Serena Xiong, J. Robin Moon, Beth Virnig, Shalini Kulasingam

## Abstract

Cervical cancer screening has reduced cervical cancer-related mortality by over 70% in countries that have achieved high coverage. However, there are significant geographic disparities in access to screening. In Ghana, although cervical cancer is the second most common cancer in women, there is no national-level cervical cancer screening program, and only 2 to 4% of eligible Ghanaian women have ever been screened for cervical cancer. This study used an exploratory, sequential mixed-methods approach to examine barriers and facilitators to cervical cancer screening from women and healthcare workers perspectives, guided by the Dynamic Sustainability Framework. Two convenience samples of 215 women and 17 healthcare personnel were recruited for this study. All participants were from one of three selected clinics (Ejisu Government Hospital, Kumasi South Hospital, and the Suntreso Government Hospital) in the Ashanti region of Ghana. Descriptive analyses were used to group the data by practice setting and ecological system. Statistical differences in means and proportions were used to evaluate women’s barriers to cervical cancer screening. Quantitative findings from the women’s survey informed qualitative, in-depth interviews with the healthcare workers and analyzed using an inductive thematic analysis. The median age of women and healthcare workers was 37.0 years and 38.0 years respectively. Most women (n=194, 90.2%) reported never having been screened. Women who had not been screened were more likely to have no college or university education. Ecologic factors identified were lack of knowledge about available services, distance to a clinic and requiring a spouse’s permission prior to scheduling. Practice setting barriers included long clinic wait times and culturally sensitive issue. The quantitative and qualitative data were integrated in the data collection stage, results, and subsequent discussion. These findings highlight the need for non-clinician-based culturally sensitive tool options for screening such as self-collected HPV tests to increase screening participation in Ghana.

## Introduction

Cervical cancer is a leading cause of cancer death among women in low-and-middle-income countries (LMIC) primarily due to a lack of access to screening (1). Ghana has a population of 10.6 million women (or approximately 63.2% of the female population) aged 15 years and older who are at risk of developing cervical cancer (2). Additionally, cervical cancer is the second most frequently diagnosed cancer in Ghanaian women with an age-standardized incidence rate of 27.4 per 100,000 women and an age-standardized mortality rate of 17.8 per 100,000 women. In contrast, despite a similar population size, Spain has age standardized cervical cancer incidence and mortality rates of 8.2 and 1.7 per 100,000 women, respectively, due to the widespread availability of screening and treatment (3).

Ghana faces many barriers in its effort to address the high incidence of cervical cancer. Currently, there is no national human papillomavirus (HPV) vaccination campaign to protect women from contracting HPV, the main cause of cervical cancer (4). As a result, both vaccination and screening rates remain low (2). The majority of women present to clinics with advanced stage cervical cancer (5). To address this, Ghana’s National Reproductive Health Policy was revised in 2014 to integrate cervical cancer screening into existing reproductive health programs such as family planning and sexually transmitted infections management services (6,7). The policy includes recommendations for screening using Visual Inspection with Acetic Acid (VIA) and Papanicolaou tests for women ages 25–45 years and cryotherapy for the treatment of precancerous cervical lesions. Despite this policy, which has been in place for nine years, only 2.4 to 4% of eligible Ghanaian women are screened for cervical cancer annually (8–11). Interviews with women in Ghana who have been diagnosed with late stage cervical cancer confirm that barriers to preventative care include high costs, a lack of knowledge, and lack of access to screening facilities (14).

Until recently, the World Health Organization (WHO) has recommended cervical cytology and VIA for cervical cancer screening in LMICs. Of the two, VIA has been more readily adopted across sub-Saharan Africa (SSA) due to the availability and low cost of acetic acid for visualizing abnormal regions, the low cost to train personnel to conduct VIA-based screening, and the ability to carry out immediate treatment if abnormal regions of the cervix are identified (referred to as ‘see and treat’). However, limitations of this approach include a low sensitivity, low reproducibility, and the need for trained personnel to conduct pelvic exams (15–18). Although cytology has an improved sensitivity compared to VIA, it also involves a pelvic exam by trained clinic staff for specimen collection, is more expensive than VIA, and requires training to interpret laboratory results (16). As a result, the WHO updated their cervical cancer screening guidelines in 2022 to primarily recommend HPV testing in conjunction with self-collected samples for women ages 30 - 50 years (19).

HPV-based screening has a significantly higher sensitivity compared to VIA and cytology for the detection of high-grade cervical lesions (20,21). HPV testing can be performed with self-collected samples, which allows patients’ privacy and, importantly, reduces reliance on clinics with trained personnel. While this approach to cervical cancer screening is appealing, its implementation in countries in SSA is not straightforward. Hurdles include determining when and where to offer screening, how to best instruct women on self-collection methods, which collection device and HPV test to use, and how to triage HPV-positive women. Deciding how to address these hurdles requires an understanding of the unique barriers faced by a population operating within a specific healthcare setting in a given country system. The Dynamic Sustainability Framework provides a structure for re-examining interventions that have already been implemented, such as cervical cancer screening in Ghana, and identifying key domains and constructs both within the practice setting itself as well as the wider ecological system that can be used to improve patient outcomes (22). To accomplish this, it’s important to understand barriers to optimizing patient outcomes from the perspective of those utilizing healthcare services as well as those providing these services. This study used the general DSF framework in combination with a mixed-methods sequential approach to identify potential barriers to cervical cancer screening from the perspectives of both women and healthcare workers.

## Materials and Methods

### Study design

Approval for this study was provided by the Committee on Human Research, Publication and Ethics, Kwame Nkrumah University of Science and Technology, Kumasi, Ghana (CHRPE/AP/043/22) and the Cancer Center Protocol Review Committee, University of Minnesota (CPRC# 2022LS045). We used aspects of the Dynamic Sustainability Framework to better understand the factors that affect the uptake of the current cervical screening practices currently implemented in Ghana. In particular, we utilized two domains, “Practice settings” and “Ecological system”, from the DSF (Fig 1) to provide a general framework for our data collection and analysis (22). In addition, we adopted an exploratory, sequential mixed-methods study design to obtain detailed information on the constructs for each of these domains. In particular, we used an initial, quantitative phase to obtain information on “other practice setting constructs” and “population characteristics” in the Ecological System domain. For this phase, we focused on Ghanaian women, overall and stratified by key characteristics. Analysis of the quantitative data informed the second, qualitative phase of the study. This second phase was designed to obtain in-depth information to better understand practice setting constraints such as staffing. In particular, we conducted key informant interviews with clinicians and laboratory workers who were directly involved in cervical cancer screening and testing at the same facilities as those used for recruitment of women eligible for screening for the quantitative data collection (Phase I). Our overall goal was to obtain information from each of these domains that could help us better understand why cervical cancer screening as an intervention may not be working well and how it can potentially be improved.

**Fig 1.**
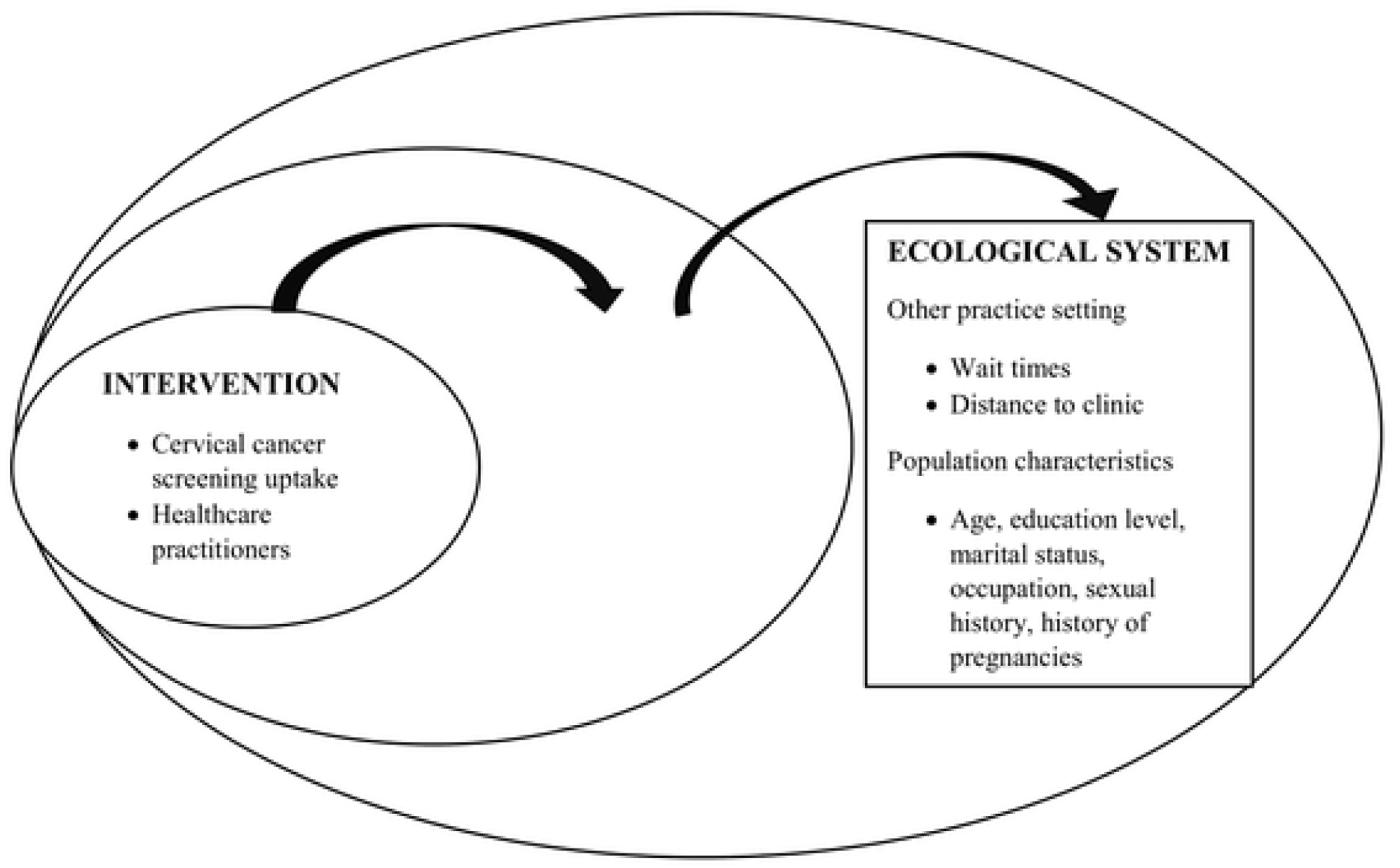
Dynamic sustainability framework for cervical cancer screening in Ghana.

### Study setting

Individuals were selected from three healthcare facilities: the Ejisu Government Hospital, Kumasi South Hospital, and the Suntreso Government Hospital. Kumasi South and Suntreso Government hospitals are urban facilities located in the city of Kumasi, Ghana’s second largest city, which has a total population of 6,630,000 individuals (23). Ejisu Government Hospital is peri-urban and is located in the municipality of Ejisu, which has a smaller population of approximately 181,000 individuals. Together, the selected facilities serve individuals from three metropolitan assemblies in the Ashanti region: Ejisu, Asokwa, and Kumasi. These areas were chosen because participation in screening is low and women served by health facilities in this region either have to wait for outreach services or travel to the few health facilities offering cervical cancer screening (24). Recruitment took place from 1^st^ August 2022 and ended on 30^th^ November 2022.

### Ecological System

#### Quantitative Data Collection (Survey)

The quantitative portion of the study examined other practice setting and population characteristics that affect women’s abilities to obtain cervical cancer screening. A survey was adapted based on a published study that evaluated the knowledge about and attitudes towards cervical cancer and cervical cancer screening in a population of 900 women living in Uganda (25). The questionnaire was drafted in English, translated into Twi and back-translated into English to check for potential errors. The survey was divided into seven sections. The first section asked for demographic information including the participant’s age, education level, marital status, occupation, average income, number of dependents, and length of living in the current district. The second and third sections asked about the participant’s knowledge of cervical cancer, cervical cancer screening and vaccination. This section included questions regarding symptoms of cervical cancer, sources of information for patient knowledge about cervical cancer and when and how often individuals should be vaccinated and/or screened for cervical cancer. The fourth section was a knowledge scale on the risk factors of cervical cancer in which participants were asked to respond ‘Yes,’ ‘No’ or ‘Don’t know’ (25). The fifth section was a belief scale that participants were asked to rank on a five-point Likert scale with ‘1’ indicating strong disagreement and ‘5’ indicating strong agreement with the statement (25). The sixth section asked about other practice setting characteristics, such as wait times and distance to clinic, that affect participation in cervical cancer screening and the final section asked about the woman’s sexual history and history of pregnancies. The questionnaire was programmed in REDCap (Research Electronic Data Capture) to allow real time data collection and uploading, and to also help with quality control and allow for efficient data processing (26,27). The cross-sectional survey was conducted at the three selected healthcare facilities as noted above. Eligible individuals were women 30 years of age and older who could undergo cervical cancer screening, lived in the catchment area of one of the three hospitals, could provide informed consent and could comprehend the survey questions either verbally or in written form. A convenience sampling approach was used to enroll women; women presenting at one of the three facilities were approached by a research assistant regarding possible interest in the study. If a woman was interested, she was invited to a private room where the research assistant provided further information and assessed participant’s eligibility. If the woman was eligible and willing to participate after disclosure of the study details, the assistant asked for consent and administered the questionnaire. All women received a compensation of GHC 30.00 for their participation in the study.

#### Analysis of quantitative data

All analyses were performed using R Statistical Software (v4.1.3; R Core Team 2021) (28). Data cleaning included evaluating missingness and non-response rates for various questions. All variables used for analysis had less than 1% of missing responses. Descriptive statistics were used to characterize population characteristics and evaluate other practice setting constraints that affect participation in cervical cancer screening. Continuous variables were assessed for median and IQR while frequencies were assessed for all categorical variables. Additionally, other practice setting constraints were stratified by education level (some college or university versus no college or university), marital status (married or living together versus single, divorced or widowed) and rural versus urban. Categorical variables were created based on prior studies examining socio-demographic variables that predict Pap-smear uptake in Ghanaian women (29). For marital status, women who were married or living with a partner were compared to women who were single, divorced, widowed, or separated. For educational status, women who had some college or university education were compared to women with secondary school education or less. Women living in rural areas were also compared to those living in urban areas, which has previously been shown to influence uptake of cervical cancer screening (30). After stratifying by demographic variables, prevalence ratios were used to assess differences in response frequency by demographic category, and two-sided t-tests were used to analyze a difference in means and differences in proportions for continuous variables. All statistical tests were deemed significant at the α=0.05 level.

#### From quantitative to qualitative methods

For the second phase, a semi-structured interview guide was developed that incorporated findings that emerged as important and were related to the Practice Setting during the first phase. Based on women’s reporting of limited accessibility to clinics, needing permission to schedule a doctor appointment, and clinic wait times of one hour or greater, interview questions explored the views of healthcare workers on health system hurdles faced by women accessing preventive care.

### Practice Setting

#### Qualitative section (Interviews)

##### Understanding barriers to cervical cancer screening from a health system perspective

To better understand how healthcare settings, affect cervical cancer screening, we interviewed healthcare workers from each of the three hospitals (the Ejisu Government Hospital, Kumasi South Hospital, and the Suntreso Government Hospital). Healthcare workers were defined as nurses/midwives, doctors or laboratory personnel who were involved in cervical cancer screening (nurses/midwives and doctors) or testing (laboratory personnel). Individuals were eligible for the study if they were 21 years of age or older, involved in cervical cancer screening, could comprehend the questionnaire verbally, and could provide informed consent. Possible participants were identified by administrators at each selected facility. Research assistants contacted each of the possible participants to review the study details and eligibility criteria. Eligible participants decided on the date, time and venue for the interview. All of the interviews were conducted in a private office within the health facility to guarantee participant comfort and privacy. Eligible individuals interested in participating were asked for consent prior to administering the interview. Total sample size was 17 and this was based on the principle of theoretical saturation, which was determined from the interviews (31). We determined we had reached data saturation when additional interviews were redundant to previously collected data (31). By the 15^th^ interview, there was no new information. We carried out two more interviews to confirm that we had reached data saturation. In total, 17 interviews were conducted. The healthcare workers’ questionnaire was split into two sections with the first section asking for demographic information such as age, gender, education level and length of employment at the current location. The second section asked about current cervical cancer screening procedures at their respective clinic and potential constraints and facilitators to accessing screening by women. This section used semi-structured interview questions and data was collected through face-to-face in-depth interviews that were audio recorded. On average, the interviews lasted 40 minutes.

#### Qualitative data analysis

Qualitative data analysis followed an inductive thematic analysis framework. The process of analysis began with verbatim transcription of both the English (n=4) and local language (Twi, n=13) audio-recorded data. For the interviews conducted in Twi, two expert translators were used in a back-back translation process. A random selection of audio data was evaluated by a bilingual research assistant to ensure accuracy in interviewing, transcribing and translation. Transcripts were imported into QSR NVivo-12 Plus for data management and coding. The ‘nodes’ function in QSR NVivo-12 was used for preliminary inductive coding (32). Intercoder agreement was 95% for codes on questions regarding barriers to cervical cancer screening and 90% agreement was observed on questions about factors that would facilitate a woman to seek cervical cancer screening. To enhance trustworthiness of the data, strategies such as prolonged engagement, peer debriefing, member checking (n=2), audit trail, reflexivity, detailed and appropriate descriptions of the methodological processes and context were employed (33) The inductive coding results were categorized according to emerging patterns and major themes around barriers to cervical cancer screening and testing. Key quotes were extracted to reflect the various themes to support the barriers.

#### Integration of the mixed-methods data

Following the structure of an exploratory, sequential mixed-methods design, the quantitative and qualitative data were integrated in the data collection stage, results and subsequent discussion. Themes and key quotes from the in-depth interviews were used to provide additional insight into health system barriers women might face in accessing cervical cancer screening.

### Ethics Approval and Consent to Participate

Approval for this study was provided by Committee on Human Research, Publication and Ethics, Kwame Nkrumah University of Science and Technology (CHRPE/AP/043/22) and the Cancer Center Protocol Review Committee, University of Minnesota (CPRC# 2022LS045). The participants received written information about the study. Formal consent was obtained by writing and participation was voluntary and anonymous.

## Results

Population characteristics for the women’s sample are presented in Table 1a and characteristics for the healthcare worker’s sample are presented in Table 1b. The median age of women was 37.0 (IQR: 32.0 - 45.5). Overall, women had varying levels of educational attainment. The majority of women were married (69.3%, n=149) and self-employed (62.8%, n=135). The median number of dependents was four. The median age of healthcare workers was 38.0 (IQR: 36.5 - 43.5). The majority of healthcare workers identified as female (78.9%, n=15) and had received a university degree (63.2%, n=12). The median length of time that healthcare workers had worked within their profession was 12.0 years (IQR: 10.0 - 15.5).

**Table 1a:** Demographics for women participants.

| Participant Characteristic (n=215) |  |
| --- | --- |
| Age* | 37.0 (32.0–45.5) |
| **Education, n (%)** |  |
| No primary school | 12 (5.6) |
| Primary school | 22 (10.2) |
| Junior high school | 78 (36.3) |
| Senior high school | 53 (24.7) |
| Some college or university | 50 (23.2) |
| **Marital Status, n (%)** |  |
| Single | 23 (10.7) |
| Married | 149 (69.3) |
| Widowed | 18 (8.4) |
| Divorced/Separated | 12 (5.6) |
| Living with a partner | 13 (6.0) |
| **Occupation, n (%)** |  |
| Agriculture/Farming | 12 (5.6) |
| Self-employed | 135 (62.8) |
| Housewife | 12 (5.6) |
| Civil/Government/Private Sector Employee | 40 (18.6) |
| Other | 16 (7.4) |
| **Dependent** |  |
| Number of Dependents* | 4.0 (3.0–5.5) |

| Length Living in District, n (%) |  |
| --- | --- |
| All of my life | 19 (8.8) |
| Less than 1 year | 22 (10.2) |
| Between 1 and 5 years | 74 (34.4) |
| More than 5 years | 100 (46.5) |

\*Age and number of dependents are reported as median (IQR)

**Table 1b:** Demographics of healthcare workers.

| Participant Characteristic | All healthcare workers (n=19) |
| --- | --- |
| Age* | 38.0 (36.5–43.5) |
| Gender, n (%) |  |
| Female | 15 (78.9) |
| Male | 4 (21.1) |
| Education, n (%) |  |
| Some college | 7 (36.8) |
| A college/university degree | 12 (63.2) |
| Employment |  |
| Length of employment in years* | 12.0 (10.0–15.5) |

\*Age and length of employment are presented in median (IQR)

Quantitative data describing other practice setting issues that women face when accessing cervical cancer screening are presented in Table 2. The median distance that women reported traveling to get to the closest healthcare facility was 5.5 kilometers (IQR: 3.1 - 10.1) and the median travel time to the clinic was 25.0 minutes (IQR: 15.0 - 30.0). One hundred and twenty-four women (57.7%) reported that they had to wait longer than one hour before being seen at the clinic. The majority of women took public transportation to get to the healthcare facility (58.1% or n=125). Aside from public transportation, women usually took either their own vehicle or a ridesharing vehicle (31.6% of n=68). Seventy-three women (34.0%) responded that they needed permission prior to scheduling a doctor’s appointment. One hundred and thirty-five women (62.3%) reported that they knew of healthcare facilities that provided cervical cancer screening; however, most women (90.2% or n=194) reported that they had never been screened for cervical cancer.

**Table 2:** Other practice setting constraints for accessing cervical cancer screening for all women (n=215)

| Factor | All Women |
| --- | --- |
| How far is it, in kilometers, to the local healthcare facility/ clinic from your home?* | 5.5 (3.1 - 10.1) |
| How much time, in minutes, does it take you to get to the clinic?* | 25.0 (15.0 - 30.0) |
| Wait longer than 60 minutes to be seen at the clinic n (%) | 124 (57.7) |
| **Primary mode of transportation, n (%)** |  |
| Car (either own or rideshare) | 68 (31.6) |
| Bicycle | 1 (.004) |
| Walking | 18 (8.4) |
| Public Transportation | 125 (58.1) |
| Needs permission to schedule a healthcare visit n (%) | 73 (34.0) |
| Knows of clinics/hospitals that offer cervical cancer screening n (%) | 80 (37.2) |
| Has never been screened for cervical cancer n (%) | 194 (90.2) |
| Has access to a mobile phone n (%) | 203 (94.4) |

\*\*Distance to the local healthcare facility and time to clinic are reported as median (IQR)

Potential constraints stratified by highest level of educational attainment, marital status and rural versus urban are presented in Table 3. When stratified by education level, women with no college or university were significantly more likely to have to wait >1 hour to be seen at a local clinic versus women with some college or university-level education (p = 0.001), and were significantly less likely to be aware of local clinics offering screening services (p = 0.05). When stratified by marital status, single, widowed, and divorced women were significantly more likely to walk to the clinic versus women who were married or living with their partners (p = 0.03). Married women were more likely to require permission to schedule a doctor appointment versus single women (p < 0.001). When comparing women living in an urban versus rural area, women living in an urban area spent, on average, a longer duration of time traveling to their local clinic.

**Table 3:** Individual-level constraints stratified by education level, marital status, and rural versus urban for all women (n = 215)

|  |  |  | Prevalence Ratio | Test Statistic | p-value |
| --- | --- | --- | --- | --- | --- |
| **Facility-level barrier** | **No College/University (n = 165), %(n)** | **Some College/University (n = 50), %(n)** |  |  |  |
| Distance (km) to local healthcare facility, mean (SD) | 8.7 (11.4) | 6.5 (6.5) |  | 1.7 | 0.09 |
| Time (minutes) to local healthcare facility, mean (SD) | 30.8 (23.2) | 27.0 (23.3) |  | 1 | 0.32 |
| >1 hour wait time at clinic %(n) | 64.8 (107) | 34.0 (17) |  | 13.7 | <0.001 |
| **Primary mode of transportation %(n)** |  |  |  |  |  |
| Car (own or rideshare) | 24.8 (41) | 54.0 (27) | Referent |  |  |
| Bicycle | 0.6 (1) | 0 | NA |  |  |
| Walking | 9.7 (16) | 4.0 (2) | 0.54 |  | 0.41 |
| Public transportation | 63.6 (105) | 40.0 (20) | 0.78 |  | 0.49 |
| Needs permission to schedule visit %(n) | 32.7 (54) | 38.0 (19) |  | 0.27 | 0.6 |
| Knows of clinics with screening %(n) | 33.3(55) | 50.0 (25) |  | 3.88 | 0.05 |
| Never been screened for cervical cancer %(n) | 90.3 (149) | 90.0 (45) |  | <0.001 | 1 |
|  |  |  | Prevalence Ratio | Test Statistic | p-value |
| **Facility-level barrier** | **Married/Living Together (n=162), n(%)** | **Single/Divorced/Widowed (n=53), n(%)** |  |  |  |
| Distance (km) to local healthcare facility, mean (SD) | 8.4 (11.1) | 7.6 (8.2) |  | 0.56 | 0.58 |
| Time (minutes) to local healthcare facility, mean (SD) | 30.0 (23.3) | 30.0 (23.4) |  | 0.002 | 1 |
| >1 hour wait time at clinic %(n) | 60.5 (98) | 49.1 (26) |  | 1.7 | 0.19 |
| **Primary mode of transportation**<br>**%(n)** |  |  |  |  |  |
| Car (own or rideshare) | 32.7 (53) | 28.3 (15) | Referent |  |  |
| Bicycle | 0 | 1.9 (1) | NA |  |  |
| Walking | 5.6 (9) | 17.0 (9) | 2.3 (1.2 - 4.3) |  | 0.03 |
| Public transportation | 59.9 (97) | 52.8 (28) | 1.0 (0.58 - 1.8) |  | 0.96 |
| Needs permission to schedule visit %(n) | 42.6 (69) | 7.5 (4) |  | 20.3 | <0.001 |
| Knows of clinics with screening %(n) | 40.1 (65) | 28.3 (15) |  | 1.9 | 0.17 |
| Never been screened for cervical cancer<br>%(n) | 88.3 (143) | 96.2 (51) |  | 2.0 | 0.15 |
|  |  |  | **Prevalence Ratio** | **Test**<br>**Statistic** | **p-value** |
| **Facility-level barrier** | **Rural (n=70)** | **Urban (n=145)** |  |  |  |
| Distance (km) to local healthcare<br>facility, mean (SD) | 9.2 (6.9) | 7.8 (11.8) |  | -1.1 | 0.26 |
| Time (minutes) to local healthcare<br>facility, mean (SD) | 24.2 (18.2) | 32.7 (24.9) |  | 2.8 | 0.005 |
| >1 hour wait time at clinic %(n) | 61.4 (43) | 55.9 (81) |  | 0.39 | 0.53 |
| **Primary mode of transportation**<br>**%(n)** |  |  |  |  |  |
| Car (own or rideshare) | 32.9 (23) | 31.0 (45) | Referent |  |  |
| Bicycle | 0 | 0.7 (1) | NA |  |  |
| Walking | 5.7 (4) | 9.7 (14) | 1.18 (0.87 - 1.59) |  | 0.37 |
| Public transportation | 57.1 (40) | 58.6 (85) | 1.03 (0.83 - 1.27) |  | 0.8 |
| Needs permission to schedule visit %(n) | 31.4 (22) | 35.2 (51) |  | 0.15 | 0.7 |
| Knows of clinics with screening %(n) | 27.1 (19) | 42.1 (61) |  | 3.9 | 0.05 |
| Never been screened for cervical cancer<br>%(n) | 87.1 (61) | 91.7 (133) |  | 0.66 | 0.42 |

### Qualitative Results

Based on the results of the quantitative analysis, two main themes related to the practice setting were further explored in these interviews: client-level constraints and health system challenges. The sub-themes that emerged through thematic analysis are presented in Table 4.

**Table 4:** Emerging practice setting relevant themes from healthcare worker interviews.

| Themes | Sub-themes |
| --- | --- |
| Client level barriers | Financial constraints and cost of treatment |
|  | Clients' non-compliance due to shyness and feelings of discomfort |
|  | Fatalistic views about the outcome of cervical cancer screening |
| Health system challenges | Infrastructural inadequacies |
|  | Logistical constraints |
|  | Low staff strength |

Healthcare workers reported that women perceived financial constraints to cervical cancer screening either through the direct cost of screening services, or the indirect cost associated with transportation to the clinic. Healthcare workers also noted that women often do not think the financial burden of screening services is worth the benefit it provides in detecting cervical cancer:

> *“Most of them complain about finances. They think that why should I use my money to go and do screening while I can use it for other pressing matters. So, it is not seen as a priority.”* (P00 2, laboratory personnel)

Healthcare workers also discussed non-compliance with screening due to feelings of discomfort around the VIA procedure. Women experienced shyness for several reasons, including fear of judgment on personal hygiene or discomfort from healthcare workers seeing them naked:

> *“Some women are shy of themselves for their fellow women to see their nakedness…this makes them non-compliant and difficult to handle. At the end of the day, you will not be able to do the screening because they will be reluctant to follow the screening procedures.”* (P006, midwife)

Fears about the results from the screening process was another theme that emerged from healthcare worker interviews. According to the key informants, their clients feared that the screening would reveal that they had the disease (cervical cancer). This fear about a positive screening result was attributed to fatalistic views held by the clients. That is, healthcare workers thought that clients feared that once they had been diagnosed as having cervical cancer, it would mean that they were going to die. Consequently, this fear made the clients non-compliant and unwilling to get screened.

> *“Fear of the unknown. You know after educating the women and giving them counseling, they will still not feel comfortable for you to screen them because of fear of the unknown. She doesn’t know what the outcome will be. The fear being diagnosed of cervical cancer and dying from the disease.”* (P00 14, midwife)

The overarching theme of practice setting constraints was further broken into three sub-themes: infrastructural inadequacies, logical constraints, and lack of staff. Healthcare workers noted that the infrastructural capacity of healthcare facilities was not adequate for providing women’s health services. For example, clinics lacked beds necessary for cervical cancer screening, or the clinic set-up did not allow for patient privacy. In clinics with a proper examination room, patients often experienced long wait times due to the availability of only one or a few rooms:

> *“We don’t have a specific room with a special bed. The room that we use is not ideal…it does not support the privacy of the client in any way. Because of that, the clients don’t feel comfortable getting screened.”* (P001, midwife)

Lastly, healthcare workers noted that inadequate staffing of clinics resulted in longer wait times. This prevents patients from returning for future visits, and patients may also advise against others wanting to use the clinic’s services:

> *“Inadequate staffing is the big issue. We are few, hence, the women wait for a long time to be attended to. They go and do not come back. Sharing such experience with other women also discourage screening uptake.”* (P0016, midwife)

## Discussion

This study illustrates the constraints that different subgroups of women in Ghana face when accessing cervical cancer screening services and highlights areas in both the ecological system and practice setting that need to be addressed to increase uptake of screening. In terms of ecological factors affecting uptake, a key constraint is lack of knowledge about facilities offering cervical cancer screening. This finding is consistent with studies conducted in Uganda (34) and Tanzania (35). The 2016 study in Uganda found that less than half of surveyed participants were aware of cervical cancer screening services (34), and the 2012 study in Tanzania found that women with a higher level of cervical cancer prevention knowledge were significantly more likely to partake in screening services (35). The implication of this finding is that individuals who intend to get screened may miss opportunities for screening due to their lack of awareness about facilities that offer screening. We also found that women with lower educational attainment were less likely to be aware of facilities that provide cervical cancer screening services. This is not surprising as higher educational attainment tends to empower women to access information including health information such as where to access cervical cancer screening services (36,37).

Contrary to previous studies that have found long travel time for cervical cancer screening among women in rural areas (38,39), we found that long travel times were more likely to be experienced by those living in urban areas. This is possibly due to higher population density, traffic congestion, or longer distances between residences and healthcare facilities in urban areas compared to rural areas. Further research is required to fully comprehend why urban dwelling women in Ghana spend long travel time in accessing cervical cancer screening services. Another key consideration is long wait times, which are a major barrier to accessing cervical cancer screening with more than half of participants (57.7%) waiting over 60 minutes to be seen at the clinic. This result is consistent with a study in which 86% of a sample of 200 Kenyan women reported long wait times as a barrier to cervical cancer screening participation (40). Similar findings were also reported in a qualitative study of 48 women conducted in Accra, Ghana (41).

In terms of population characteristics, our study also highlights the fact that women with lower educational attainment were more likely to experience longer wait times at local clinics. A plausible explanation for this finding could be that women with higher education are more likely to access cervical cancer screening services in private healthcare facilities where there is less congestion and faster service delivery. This study did find that women with higher educational attainment received, on average, higher monthly incomes, which increases access to more expensive, private clinics. Women with lower educational attainment may be more likely to access cervical cancer screening services at public healthcare facilities where there is a high volume of patients, long queues, and fewer staff available for screening (42). These factors slow service delivery consistent with our qualitative results from interviews with healthcare workers who asserted that the low staffing in their facilities resulted in longer waiting times that discouraged women from accessing cervical cancer screening services. Another perspective is that lower educational attainment may be a proxy for socioeconomic status and lower information attainment. This is in the sense that women with lower levels of education often face limited access to economic opportunities, resulting in lower income levels and financial resources. This economic disadvantage can directly impact their ability to access healthcare services, including cervical cancer screening, due to financial constraints such as transportation costs or inability to afford screening fees.

Consistent with another study conducted in Pakistan (36), we found that needing permission to schedule appointments was a barrier to women’s access to cervical cancer screening. The study revealed that married women were more likely to face the challenge of needing permission before they could schedule a visit. This finding may be explained by existing patriarchal and gender norms that place the male as the decision-maker in the household (43,44). Consequently, the autonomy of women to make decisions regarding their healthcare is significantly reduced, especially among married women. Ghanaian traditional culture also encourages women to ensure that their nakedness is only seen by their husbands or partners. Given that the cervix is examined during cervical cancer screening, married women may feel uncomfortable undergoing this process (12). Those willing to undergo screening would then require their partners to consent and grant permission before they can allow another person to see their ‘nakedness.’ This assertion was corroborated by the accounts of the healthcare workers regarding shyness and discomfort women feel in relation to cervical cancer screening. These results suggest that other forms of screening such as HPV test-based screening through self-collected samples could increase screening uptake by mitigating patient shyness and the need for permission. Self-collection does not require the assistance of healthcare providers, which increases the privacy of the screening procedure. Previous studies have shown that women feel comfortable and confident with this type of screening (45). Further, this type of screening may also increase screening by circumventing the need for clinic-based screening.

In conclusion, this study finds that a large proportion of Ghanaian women have never been screened for cervical cancer. Both the quantitative and qualitative data highlight major barriers that women face in access to cervical cancer screening. The women’s survey found that long clinic wait times may impact screening uptake. This was especially true for women with lower educational attainment (i.e. those with no college or university education). Additionally, there was an overall lack of knowledge regarding where to obtain screening services, consistent with other studies previous findings. In-depth interviews with healthcare workers confirmed that healthcare facilities are not adequately staffed, resulting in longer patient wait times. Healthcare workers also noted culturally sensitive issue as a barrier to screening uptake. This expanded upon the quantitative data, which found that over one-third of surveyed women (and over two-thirds of surveyed married women) needed permission prior to scheduling a doctor appointment. Moving forward, the use of self-collected samples for HPV testing could mitigate barriers such as time spent traveling to and waiting at a healthcare clinic. It might also provide a more culturally sensitive tool to address issues of cultural norms because HPV sample self-collection does not require the assistance of healthcare providers. Future studies should assess the feasibility of implementing self-collected HPV samples as a possible method for cervical cancer screening.

## Data Availability

The datasets generated and/or analyzed during the current study are not publicly available due to ethical reasons but are available from the corresponding author on reasonable request.

## Declarations

### Author’s Contributions

- ABBM: Conceptualization, methodology, data curation, formal analysis, investigation, project administration, writing – original draft, writing – review & editing
- TOK: Conceptualization, methodology, investigation, data curation, writing – review & editing
- EA: Methodology, investigation, writing – review & editing
- JA: Methodology, investigation, writing – review & editing
- MN: Methodology, data curation, formal analysis, investigation, writing – original draft, writing – review & editing
- JO: Data curation, formal analysis, writing – original draft, writing – review & editing
- KOB: Methodology, investigation, writing – review & editing
- FMEE: Methodology, investigation, writing – review & editing
- SX: Methodology, writing – review & editing
- RM: Methodology, writing – review & editing
- BV: Conceptualization, methodology, data curation, investigation, writing – review & editing
- SK: Conceptualization, methodology, data curation, formal analysis, investigation, project administration, writing – original draft, writing – review & editing

## Acknowledgments

We are grateful to all participants who shared their experiences in this study and funder-Institute of Global Cancer Prevention Research (IGCPR). Masonic Cancer Center, University of Minnesota.

